# A Quantitative Framework for EHR Cohort Refinement

**DOI:** 10.64898/2026.04.27.26351837

**Authors:** Nattanit Songthangtham, Gyorgy Simon, Steven G. Johnson

## Abstract

Electronic Health Record (EHR) based research depends on accurate cohort definitions, yet current workflows offer little early insight into cohort quality and rely heavily on slow, ad hoc manual review. We developed a quantitative, iterative framework that integrates model-guided case selection with sequential statistical testing to provide an early, data-driven signal of cohort accuracy. Using an OMOP-standardized dataset and the PCORnet Type 2 Diabetes phenotype as the gold-standard proxy, we evaluated eight sampling strategies across starting review sizes from 30 to 2,000 cases. At each iteration, the framework retrained an internal logistic regression model, selected cases for review, and applied a Bonferroni-adjusted Agresti–Coull upper bound to assess whether the cohort met a pre-specified accuracy threshold. Across 48 simulation conditions, starting sample size strongly shaped iteration count and total review burden, and adaptive sampling strategies consistently required fewer reviewed cases than fixed-batch methods. These findings demonstrate a reproducible, statistically grounded approach for refining EHR cohort definitions while reducing manual review effort.

## 1. Introduction

Electronic Health Records (EHRs) now support a wide range of biomedical research activities. They enable large-scale retrospective studies, pragmatic trial emulations, and real-world evidence generation^1,2^. Two distinct roles are usually involved when EHRs are used in clinical research. Researchers, typically clinicians or domain experts, originate the research question and specify the conceptual cohort definition through data request forms. Analysts then translate these conceptual definitions into executable queries and extract data from clinical warehouses ^3^. Prior work shows that researchers often provide criteria that are ambiguous, underspecified, or difficult to operationalize, requiring analysts to interpret clinical intent and resolve inconsistencies before data can be retrieved ^3–5^. Because clinical concepts rarely map cleanly to structured fields, initial cohort definitions are frequently broad or imperfect, and translating clinical ideas into computable logic remains a central challenge in EHR-based research^6,7^.

Despite the centrality of EHR data in clinical research, current cohort-construction workflows provide researchers with little early insight into whether a cohort is usable, improvable, or fundamentally misaligned with the intended clinical concept. Instead, cohort refinement unfolds through slow, iterative cycles in which analysts re-execute revised definitions and researchers reassess the resulting cases. Prior studies describe this process as a major bottleneck. The back-and-forth clarification and re-extraction often consume more time than the technical retrieval itself and routinely dominate project timelines^2,8^. These inefficiencies have measurable consequences. Misclassification bias, inconsistent phenotype definitions, and poor reproducibility remain pervasive across EHR-based studies^7,9–12^. A study by Ostropolets et al.^13^ shows that even when experienced teams share code, agreement with expert-validated cohorts can be strikingly low, with a median overlap of only 9.4%^13^. Yet manual review, the primary mechanism for detecting these issues, is neither standardized nor optimized; cases are often selected ad hoc, and review typically occurs only after substantial effort has already been invested. As a result, researchers frequently discover too late that a cohort is unfit, leading to wasted time, wasted effort, and missed opportunities for timely scientific discovery.

This lack of standardization is frequently attributed to ad hoc workflows and the divide between clinicians and data scientists, in which clinicians provide broad conceptual definitions that analysts must translate into operational criteria^14,15^. Additional structural barriers such as variability in EHR documentation, inconsistent use of coding systems, and interoperability challenges across data models, can further complicate efforts to construct reproducible cohorts^12,16,17^. These issues are compounded by the complexity of query mediation and the broader challenges associated with re-using EHR data for research^3–5^. Recent work further emphasizes that even when phenotype algorithms exist, their intended population and performance characteristics are often underspecified. The PhenoFiT algorithm, from a study by Wiley et al.,^18^ shows how hidden design choices and site-specific data constraints can cause algorithms to identify non-equivalent populations or lose performance when transferred across settings. The authors emphasized the need for early and systematic verification of cohort quality. Because initial cohort definitions influence all downstream analyses, there is a critical need for systematic cohort verification to identify quality issues before the final analysis phase.

To address these challenges, we propose a structured, quantitative framework that helps refine cohort definitions through an iterative process rather than a linear sequence of data pulls. The framework utilizes intelligent case-selection logic integrated with a statistically grounded stopping rule to provide a real-time signal of cohort quality. By requiring an initial round of manual review on a systematically selected sample, the framework generates a rapid estimate of the cohort error rate and its associated uncertainty. This approach allows the refinement process to be guided by quantitative metrics, enabling researchers to determine precisely when a cohort has reached a predefined quality threshold.

We evaluate this framework through a series of simulations to characterize its performance across different operational parameters. We test eight distinct case-selection strategies, including both fixed-size and adaptive-size sampling methods, to quantify their efficiency in terms of total cases reviewed and total iterations required for convergence. To ensure the framework’s robustness as a generalizable alternative method, we evaluate it across varying levels of data noise and reviewer error and across multiple starting clinical definitions. Our goal is to demonstrate that this framework provides a principled, evidence-based signal for measuring cohort fitness, significantly reducing manual effort while maintaining rigorous statistical certainty. We do not aim to identify an optimal evaluation strategy; rather, we show that the framework can quantify cohort accuracy across diverse starting definitions and can serve as a foundation for discovering optimal strategies in future work.

## 2. Methods

Manual record review remains the standard approach for determining whether an extracted dataset accurately represents a clinical cohort. However, reviewing every patient record is labor-intensive and time-consuming. To address this limitation, we developed an iterative framework to help make this process more efficient and effective. The framework aims to reduce both the total case load for manual review and the number of iterations required. The framework outputs a quantitative metric that provides a statistical guarantee that represents how well the cohort aligns with the intended target population.

We utilized an EHR dataset from Fairview Health Services, standardized to the OMOP Common Data Model (v5.3)^19^. To demonstrate the framework’s capabilities, we need a “gold-standard” intended cohort and a starting point. Type 2 Diabetes Mellitus (T2DM) served as the target condition, and the PCORnet phenotype definition was used as the gold-standard proxy for determining labels. All simulation runs were initialized with a starting definition based on the ICD-10 diagnostic code E11.9, which we selected because it provides the simplest and most direct way to specify our example T2DM cohort for initialization. All simulations were implemented in Python.

### 2.1 The Framework

The framework formalizes the interaction between clinical expertise and data analysis by structuring the cohort refining process as an iterative, data-driven workflow. Throughout the process, the framework provides continuous, transparent feedback through quantifiable metrics, allowing the researcher to monitor progress and refine the cohort definition over time. The workflow operates as a feedback loop and is supported by two distinct models. A rule-based cohort definition supplied by the researcher, which is the object of refinement, and an internal logistic regression model that is updated at each iteration and used solely to guide case selection. Together, these components enable the framework to systematically identify errors, prioritize informative cases, and converge toward a more accurate and reliable cohort definition. The framework operates as a feedback loop depicted in Figure 1:

**Figure 1.**
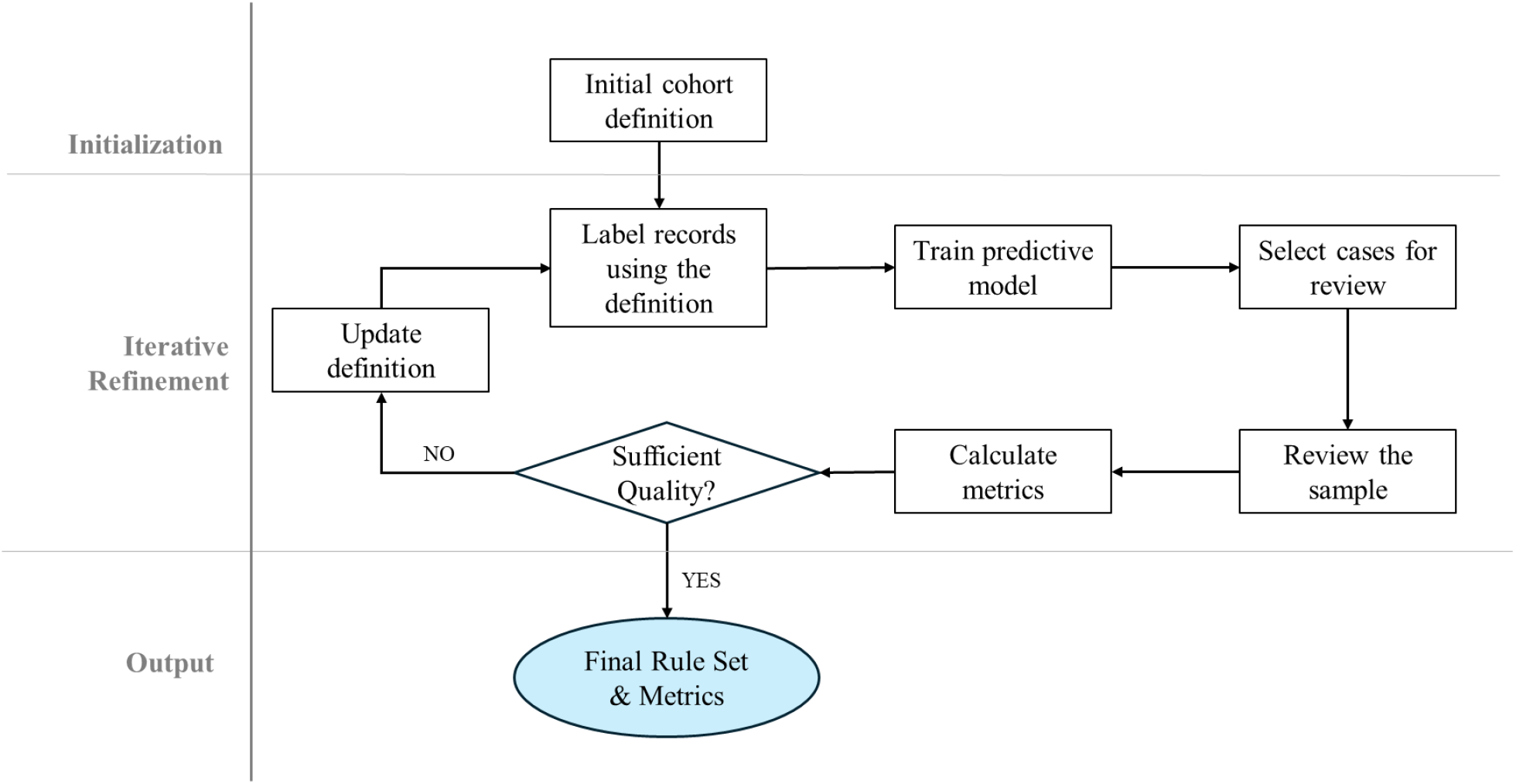
An Overview of the Framework.

#### 2.1.1 Initialization

The iterative process begins with an initial cohort definition, developed using the researcher’s conceptual “best guess” logic, any relevant phenotype library elements, and the available population dataset. A domain expert specifies the intended cohort in clinical or operational terms, and an analyst translates this logic into a computable Boolean rule set using structured data elements. The rule set gets applied to the population dataset to generate the initial binary indicator of cohort membership. This process anchors the first iteration.

#### 2.1.2 Iterative Refinement

##### Label Records Using the Current Definition

Once the initial cohort definition has been applied to the dataset, the framework enters the main loop. At each iteration, the framework labels all records according to the current rule and uses these labels to guide case selection for manual review.

##### Train Predictive Model

Case selection is supported by an internal predictive model that is updated at every iteration. The logistic regression was used as an internal model for this study. In the first iteration, the model is trained using reviewer-assigned labels from a simple random sample drawn from the starting cohort. In subsequent iterations, the model is retrained using all accumulated reviewer labels as the outcome and the binary data elements referenced in the rule as predictors. The fitted model generates predicted probabilities for all unreviewed cases, indicating how closely each case aligns with the current definition.

##### Select Cases for Review

Case selection is supported by an internal logistic regression model that is updated at every iteration and serves as a refinement tool for identifying cases that may not align with the rule. The predicted probabilities calculated from the model are used to select the next set of cases for the researcher to review. Once a case has been reviewed, it is removed from the pool of eligible cases and is never reviewed again. We wanted the framework to be able to test the effectiveness of different sampling strategies. To demonstrate this, we developed eight sampling strategies, organized into fixed-batch and adaptive-sizing families.

Fixed-Batch Strategies: Fixed-batch strategies use a constant number of cases per iteration and differ only in how they sample across the distribution of predicted probabilities from the logistic model.

1. Fixed Random: Each batch is drawn uniformly at random.
2. Fixed 25–50–25 Buckets: Cases are partitioned into three probability strata. The upper stratum consists of cases that fall within the top 20% of the predicted probability distribution. The bottom stratum holds cases that fall within the bottom 20% of the predicted probability distribution. The middle stratum comprises the rest of the cases. The 25-50-25 selection logic reflects the proportion of cases being selected from each stratum, where 25% of the selected cases come from the upper stratum, 25% from the lower stratum, and 50% from the middle stratum to balance exploration of certain and uncertain regions.
3. Fixed 25–50–25 Buckets + Random: Sampling follows the 25–50–25 structure until a predefined threshold for convergence is reached, after which the strategy reverts to random sampling. The threshold of convergence represents the point where failure rate reaches a desired value.
4. Fixed Middle Buckets + Random. Sampling is restricted to the middle stratum to prioritize cases most likely to refine the decision boundary. When this stratum is depleted or the threshold for convergence is reached, the strategy defaults to random sampling.

Adaptive-Sizing Strategies: Strategies 5–8 extend the four fixed-batch structures by replacing the static batch size with a dynamically computed quantity. In each iteration, the required number of cases is recalculated based on the current observed failure rate and the statistical precision needed to further constrain the Agresti–Coull upper confidence bound toward the convergence threshold.

The adaptive batch size corresponds to the minimum number of cases needed to identify at least two additional errors. Adaptive sizing is applied to the same four structural sampling logics:

5 Adaptive Random
6 Adaptive 25–50–25 Buckets
7 Adaptive 25–50–25 Buckets + Random
8 Adaptive Middle Buckets + Random

##### Review the Sample

Selected cases are presented to the reviewer for manual review. At least one round of review is required as the framework cannot estimate error rates or train the logistic regression model without any reviewer-assigned labels. The reviewer determines whether each case truly belongs to the target cohort as defined by the researcher’s intended population. Under standard use, this would involve a domain expert applying clinical judgment and available documentation, and the framework assumes these reviewer-assigned labels are correct. We also provide an example of how the framework works if a reviewer is not always correct. In this study, the review process is simulated using a “simulated researcher” that provides a gold-standard proxy designed to emulate a researcher’s decisions consistently and reproducibly.

During review, each case is shown alongside the current cohort definition. The reviewer assigns a binary label indicating whether the case should be included in the cohort. This label is compared with the case’s classification under the current rule to produce a status category that reflects agreement or disagreement between the rule and the reviewer. These status categories identify where the rule is performing correctly and where it is failing, and they provide the information needed for potential rule revision in the next step of the framework. Once a case has been reviewed, its label is fixed and never updated. The output of this step is the expanded set of reviewed cases, which feeds directly into updating the logistic regression model, estimating error rates, and determining whether the cohort definition should be revised

##### Calculate Metrics

Following each round of manual review, the framework calculates an error rate and evaluates whether the refinement process should continue. The framework uses the calculated error rate at the current iteration to compute the upper bound of the Agresti–Coull confidence interval, which provides a conservative estimate of the rule’s error rate based on all cases reviewed to date. Once this upper bound is obtained, the reviewer is presented with the current error estimate and its statistical guarantee. The error estimate represents a failure rate, reflecting the proportion of reviewed cases that are incorrect based on the rule set. The statistical guarantee is presented using the upper bound of the confidence interval of the error estimate, representing the highest possible value of the true failure rate.

The Agresti–Coull interval is used in this framework because it provides a stable and conservative estimate of a binomial proportion, particularly in early iterations when the number of reviewed cases may be small. Because the stopping rule is evaluated repeatedly across iterations, each evaluation constitutes a formal statistical claim. To maintain control over the family-wise error rate under repeated testing, a Bonferroni correction is applied. The per-iteration significance level is updated to alpha=0.05/k, where k denotes the cumulative number of evaluations performed to date, and the corresponding z-score is recalculated at each iteration before computing the updated upper bound.

##### Decision Point: Sufficient Quality

Once the failure rate and its upper confidence bound are computed, the reviewer decides whether refinement should continue. If the current rule is deemed sufficiently accurate, the process terminates and the rule is retained along with its estimated error rate and statistical guarantee. If further improvement is warranted, the framework proceeds to the rule-revision step, where the researcher updates the cohort definition based on the errors identified during review. Any modification to the rule reshapes the pool of unreviewed cases and provides new information for retraining the logistic regression model in the next iteration.

##### Optional: Update Definition

If refinement continues, the framework proceeds to rule revision. The framework prompts the researcher to update the cohort definition to correct the errors identified during review. Any modification to the rule reshapes the pool of unreviewed cases and provides new information for retraining the logistic regression model.

To generate candidate revisions, the simulated researcher algorithm applies two internal procedures that analyze the reviewed cases and propose modifications targeted to the observed errors. One procedure examines false positives and identifies conditions that, if added, would tighten the rule and reduce inappropriate inclusions. The other examines false negatives and identifies conditions that would expand the rule to recover missed cases. Each proposed modification is expressed as a candidate rule and converted into disjunctive normal form (DNF) to ensure a consistent representation. Before further evaluation, the framework filters out any candidate rules that were selected in previous iterations to prevent cycling through the same revisions.

Among the remaining candidates, the framework identifies the rule or rules that minimize the error rate associated with each proposal, reflecting the expected reduction in misclassification if the candidate were adopted. The framework selects the candidate that yields the greatest improvement while respecting this constraint. If no viable candidates remain after filtering, or if all reviewed cases are correctly classified, the framework concludes that no revision is needed at the current iteration. Once a revised rule is selected, it becomes the new working definition for the next iteration. The framework then returns to case selection, applying the updated rule to the full dataset as the iterative refinement process continues.

#### 2.1.3 Output

At the end of the iterative refinement process, the framework returns the finalized cohort definition, the associated performance metrics, and the labeled dataset generated during review. The accompanying metrics include the estimated failure rate and the upper bound of the Agresti–Coull confidence interval at the stopping iteration, providing a statistical guarantee on the rule’s accuracy.

### 2.2 The Simulated Researcher

For this study, the manual review process is simulated rather than performed by human reviewers so that we can quickly evaluate many scenarios. The simulated researcher provides labels and applies decisions consistent with the intended phenotype. For example, we use the PCORnet^20^ Type 2 Diabetes phenotype definition as an example of what the researcher may intend. This means that all manual review and rule-revision decisions are made against the PCORnet definition acting as the gold-standard proxy. This simulation enables the framework to be evaluated under controlled and reproducible conditions.

During each round of manual review, the simulated researcher assigns each selected case a binary determination of whether it meets the cohort definition. To ensure logical coherence and efficiency, all cohort definitions generated during the revision step are simplified into Disjunctive Normal Form (DNF). For example, a researcher-defined rule may specify that a patient meets the Type 2 diabetes cohort definition if they satisfy any two of the following conditions: (i) HbA1c ≥ 6.5%, (ii) an active prescription for a glucose-lowering medication, or (iii) abnormal glucose laboratory values such as fasting plasma glucose ≥ 126 mg/dL or random plasma glucose ≥ 200 mg/dL. When expressed in DNF, this two-of-three rule expands into the following clauses: *A1cHigh & OnMedication* | *A1cHigh & FastingGlucoseHigh* | *A1cHigh & RandomGlucoseHigh* | *OnMedication & FastingGlucoseHigh* | *OnMedication & RandomGlucoseHigh*. Each clause represents a distinct pathway by which a case can satisfy the cohort definition, ensuring that the logical structure is explicit, unambiguous, and directly evaluable by the system.

Rule revision is initiated only when at least one reviewed case is determined to be either a false positive or a false negative. If all reviewed cases align with the existing definition, no revision is performed for that iteration. For this study, the simulated researcher is permitted to make one rule correction per iteration.

When rule revision is required, the simulated researcher determines how the cohort definition should be modified to correct the observed errors. The process proceeds as follows:

- Start from the current rule: The existing cohort definition provides the baseline for generating candidate revisions.
- Enumerate all candidate rule modifications: Candidate rules are generated under the assumption that the researcher seeks to minimize the overall error rate across the entire cohort. Error calculations incorporate both newly reviewed and previously adjudicated cases to ensure that revisions generalize to the full dataset.
- Apply revision operators based on error type.
  - Extension (&) to correct false positives by adding a restrictive condition.
  - Exclusion (&∼) to correct false positives by removing or negating a condition.
  - Addition (|) to correct false negatives by adding an inclusive condition.
- Select the optimal candidate rule: The simulated researcher selects the rule that yields the lowest error rate across the labeled sample. If multiple candidate rules achieve the same error rate, the rule with the fewest conditions is chosen.

### 2.3 Framework Evaluation

We next evaluated the framework’s behavior under controlled conditions. The goal of this evaluation is to characterize how the system operates across a range of parameter settings. Each simulation run is defined by two user-specified inputs: the case-selection method and the starting batch size. All manual reviews during these runs is performed by the simulated researcher described in the previous section, ensuring consistent, gold-standard adjudication across all conditions. The starting batch size determines the number of cases reviewed in the first iteration and serves as a tunable parameter that influences both the number of iterations and the total cases reviewed. In this study, we evaluated starting batch sizes of 30, 50, 100, 500, 1000, and 2000, and for each case-selection method we ran the framework across all starting sizes. All simulations used a pre-specified convergence threshold of 1% for the confidence interval upper bound or when the researcher decides to terminate the process.

#### 2.3.1 Performance Metrics

Framework performance was evaluated by examining how each case-selection method progressed through the refinement process. Two primary metrics were used to characterize both the operational burden and the speed of convergence.

- Total cases reviewed: Total cases reviewed represent the cumulative number of cases manually reviewed across all iterations. This metric reflects the overall review effort required by each selection method, with lower values indicating greater efficiency.
- Iterations: Iterations represent the number of discrete refinement cycles completed before the framework terminates. This metric captures how quickly each selection method progressed through the framework, with fewer iterations indicating faster convergence.

#### 2.3.2 Error Injection

To assess how the framework performs under controlled error conditions, we introduced two forms of injected error. Data error was applied before sampling by randomly flipping the underlying binary label for a specified proportion of the dataset. Researcher error was independently applied during manual review by randomly flipping the reviewer’s adjudicated labels. All injected errors were completely random, and all other components of the framework remained unchanged.

## 3 Results

### 3.1 Overview of Iteration and Review Burden Across Methods

Table 1 summarizes the number of iterations and total cases reviewed across all combinations of case-selection methods and starting batch sizes. The columns show the total number of iterations and total cases reviewed required for either the failure rate and the upper bound of the confidence interval to reach 1% or less. For this simulation, this marks the point where the framework terminates.

**Table 1.** Iterations and Total Cases Reviewed Across Case-Selection Methods and Starting Batch Sizes.

| Selection Methods | Starting Review Size | Failure Rate $\leq 1\%$ | | Upper Bound $\leq 1\%$ | |
| --- | --- | --- | --- | --- | --- |
|  |  | Total Iterations | Total Cases Reviewed | Total Iterations | Total Cases Reviewed |
| Fixed Random | 30 | 15 | 450 | 34 | 1020 |
| Fixed Random | 50 | 8 | 400 | 19 | 950 |
| Fixed Random | 100 | 5 | 500 | 8 | 800 |
| Fixed Random | 500 | 3 | 1500 | 4 | 2000 |
| Fixed Random | 1000 | 3 | 3000 | 4 | 4000 |
| Fixed Random | 2000 | 3 | 6000 | 4 | 8000 |
| Fixed 25-50-25 Buckets | 30 | 15 | 450 | 34 | 1020 |
| Fixed 25-50-25 Buckets | 50 | 8 | 400 | 18 | 900 |
| Fixed 25-50-25 Buckets | 100 | 6 | 600 | 8 | 800 |
| Fixed 25-50-25 Buckets | 500 | 6 | 3000 | 6 | 3000 |
| Fixed 25-50-25 Buckets | 1000 | 6 | 6000 | 6 | 6000 |
| Fixed 25-50-25 Buckets | 2000 | 5 | 10000 | 6 | 12000 |
| Fixed 25-50-25 Buckets-Random | 30 | 15 | 450 | 34 | 1020 |
| Fixed 25-50-25 Buckets-Random | 50 | 8 | 400 | 18 | 900 |
| Fixed 25-50-25 Buckets-Random | 100 | 6 | 600 | 8 | 800 |
| Fixed 25-50-25 Buckets-Random | 500 | 6 | 3000 | 6 | 3000 |
| Fixed 25-50-25 Buckets-Random | 1000 | 6 | 6000 | 6 | 6000 |
| Fixed 25-50-25 Buckets-Random | 2000 | 5 | 10000 | 6 | 12000 |
| Middle Buckets-Random | 30 | 15 | 450 | 34 | 1020 |
| Middle Buckets-Random | 50 | 8 | 400 | 18 | 900 |
| Middle Buckets-Random | 100 | 6 | 600 | 8 | 800 |
| Middle Buckets-Random | 500 | 7 | 3500 | 7 | 3500 |
| Middle Buckets-Random | 1000 | 5 | 5000 | 6 | 6000 |
| Middle Buckets-Random | 2000 | 6 | 12000 | 6 | 12000 |
| Adaptive Random | 30 | 15 | 449 | 34 | 1019 |
| Adaptive Random | 50 | 8 | 389 | 19 | 939 |
| Adaptive Random | 100 | 6 | 530 | 9 | 830 |
| Adaptive Random | 500 | 3 | 600 | 5 | 1560 |
| Adaptive Random | 1000 | 3 | 1084 | 4 | 2044 |
| Adaptive Random | 2000 | 3 | 2070 | 4 | 3330 |
| Adaptive 25-50-25 Buckets | 30 | 15 | 447 | 34 | 1017 |
| Adaptive 25-50-25 Buckets | 50 | 9 | 412 | 19 | 912 |
| Adaptive 25-50-25 Buckets | 100 | 7 | 549 | 10 | 849 |
| Adaptive 25-50-25 Buckets | 500 | 6 | 1338 | 6 | 1338 |
| Adaptive 25-50-25 Buckets | 1000 | 6 | 1542 | 6 | 1542 |
| Adaptive 25-50-25 Buckets | 2000 | 3 | 2067 | 5 | 3616 |
| Adaptive 25-50-25 Buckets-Random | 30 | 15 | 448 | 34 | 1018 |
| Adaptive 25-50-25 Buckets-Random | 50 | 9 | 419 | 20 | 969 |
| Adaptive 25-50-25 Buckets-Random | 100 | 7 | 532 | 10 | 832 |
| Adaptive 25-50-25 Buckets-Random | 500 | 6 | 1338 | 6 | 1338 |
| Adaptive 25-50-25 Buckets-Random | 1000 | 6 | 1542 | 6 | 1542 |
| Adaptive 25-50-25 Buckets-Random | 2000 | 3 | 2067 | 5 | 3616 |
| Adaptive Middle Buckets-Random | 30 | 15 | 435 | 34 | 1005 |
| Adaptive Middle Buckets-Random | 50 | 10 | 432 | 20 | 932 |
| Adaptive Middle Buckets-Random | 100 | 6 | 410 | 10 | 802 |
| Adaptive Middle Buckets-Random | 500 | 6 | 1197 | 6 | 1197 |
| Adaptive Middle Buckets-Random | 1000 | 5 | 1878 | 5 | 1879 |
| Adaptive Middle Buckets-Random | 2000 | 5 | 3350 | 6 | 3353 |

Across methods, the starting batch size exerted a consistent and substantial influence on both outcomes. Starting sizes at 100 or lower required more iterations to reach the convergence threshold, whereas larger starting sizes reduced the number of iterations but increased the total number of cases reviewed. This pattern is consistent across fixed and adaptive strategies, indicating that the starting batch size functions as a primary determinant of the overall review burden.

### 4.2 Visualizing Iteration and Review Trade-offs

Figure 2 visually illustrates iteration and review volume trade-off by plotting iteration counts (left panel) and total cases reviewed (right panel) across all methods and starting sizes. The figure highlights the central trade-off observed in Table 1. Methods that converge in fewer iterations often require substantially more total review, whereas methods with more refinement cycles frequently achieve lower cumulative burden. For starting batch sizes that are 100 or less, there is no noticeable difference between the fixed and adaptive strategies. However, at the starting batch sizes of 500 and above, there is a clear contrast in total cases reviewed between the fixed and the adaptive methods, while the number of total iterations remains consistent. This shows that the adaptive methods outperform the fixed methods much more clearly when the starting batch size is large.

**Figure 2.**
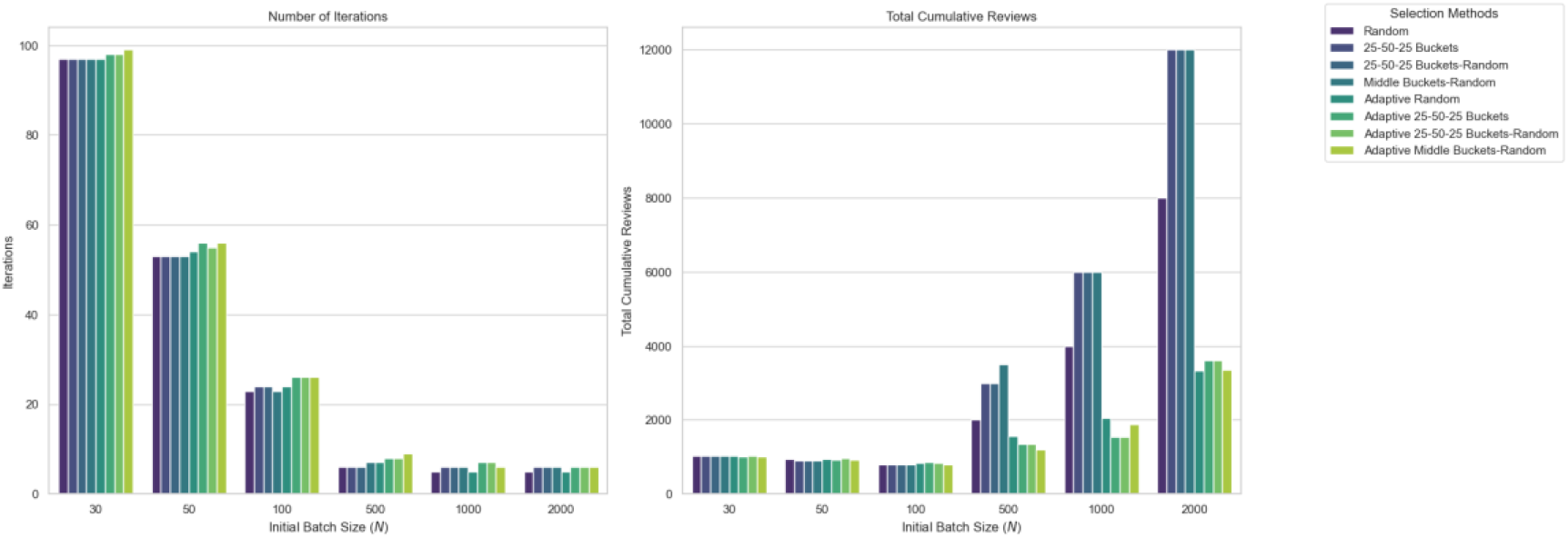
Number of Iterations and Total Cases Required to Reach Stopping Threshold Across Selection Strategies

### 4.3 Joint Relationship Between Iterations and Total Review Volume

Figure 3 further summarizes the trade-off patterns by plotting total iterations against total cases reviewed for every selection-method and starting-batch-size combination. Smaller starting sizes cluster toward the lower portion of the plot and spread widely along the horizontal axis, reflecting the higher number of iterations required when the initial sample is small. In contrast, larger starting sizes appear as larger bubbles positioned higher on the y-axis, indicating substantially greater total review volume despite requiring fewer iterations.

**Figure 3.**
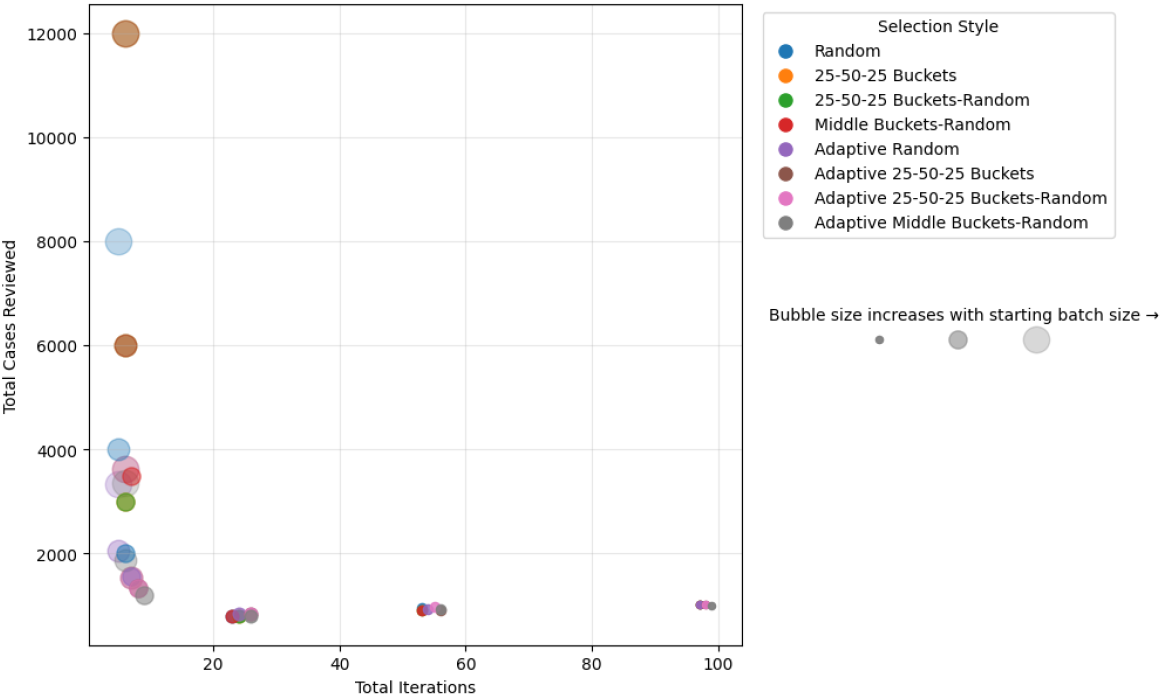
Total Iterations vs. Total Cases Reviewed for All Selection-Method and Starting-Batch-Size Combinations

### 4.4 Identifying Efficient Strategy–Size Combinations

Figure 4 isolates the lower-left quadrant of Figure 3 to highlight the better-performing combinations. The circles in the figure represent fixed selection methods where diamonds represent adaptive methods. The colors represent the different starting batch sizes. For this study, the better-performing methods are those that achieve both relatively few iterations and relatively low total review volume. Most combinations in this region fall between 5 and 10 iterations and approximately 1,000 to 2,000 total cases reviewed. Adaptive strategies dominate this space, especially Adaptive Random and Adaptive Middle Buckets–Random at starting sizes of 500 and 1000. One notable exception is the Middle Buckets–Random strategy at the starting batch size of 100, which requires 8 iterations despite a modest total review burden. This outlier underscores that low total review volume does not necessarily imply rapid convergence and that iteration count and review burden must be evaluated jointly.

**Figure 4.**
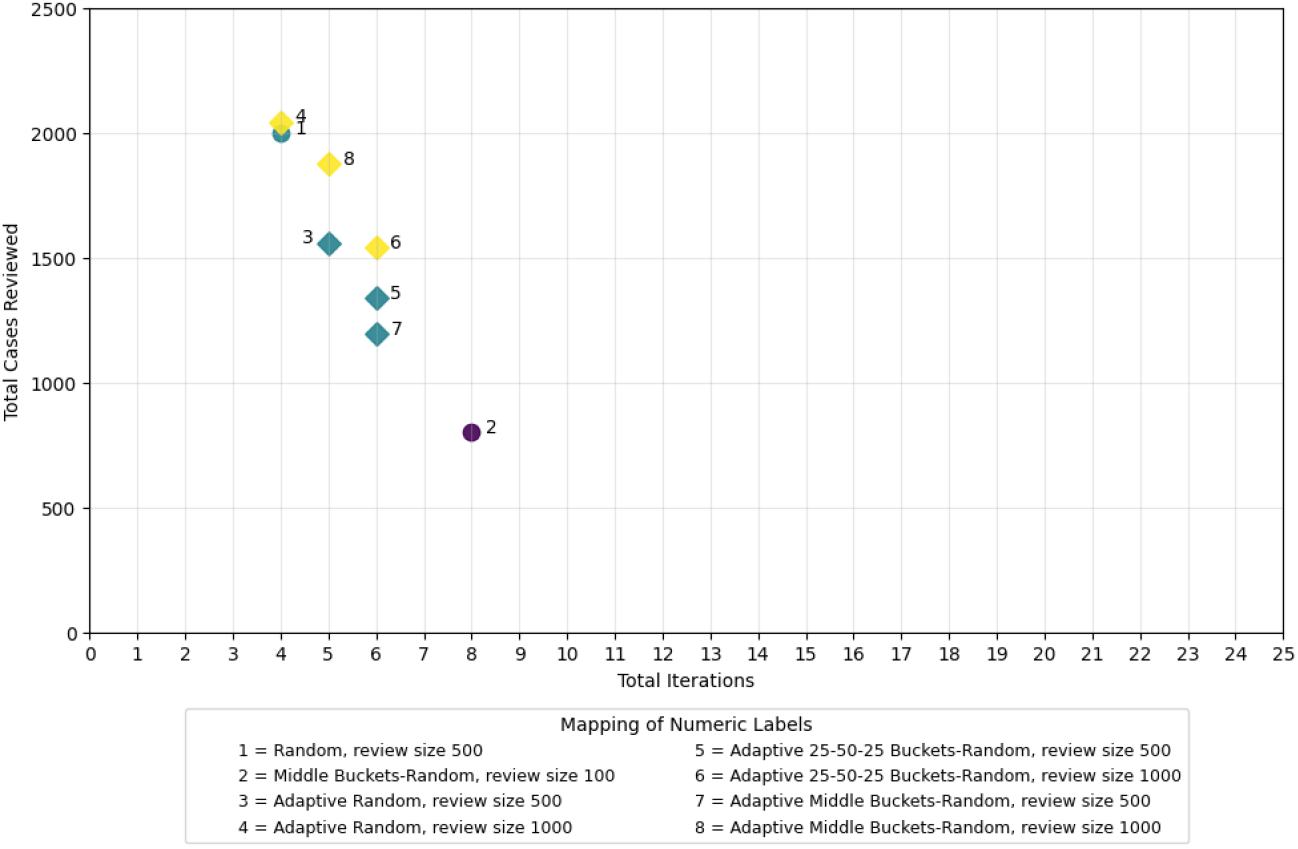
Filtered View of Better Performing Selection-Method and Starting-Batch-Size Combinations

### 3.2 Stress Testing and Robustness to Error

To evaluate the framework’s robustness to different types of error, we purposefully introduce errors into the underlying data and separately evaluate how researcher review errors impact the framework. First, we examine the effects of errors in underlying data quality. We stress-tested the selection methodologies using a fixed starting batch size of 500 cases and 4 better-performing methods from Figure 4 to ensure clarity and comparability across conditions. Figure 5 plots the failure-rate trajectories across eight iterations for injected data-error levels of 0.5%, 1.0%, 2.0%, and 5.0% where the researcher’s error is held constant at 0%. Across all error levels, the trajectories exhibit a smooth, monotonic decline, and the 95% confidence upper bound ultimately falls below the 1.0% certification threshold for every method–error combination when the framework terminates.

**Figure 5.**
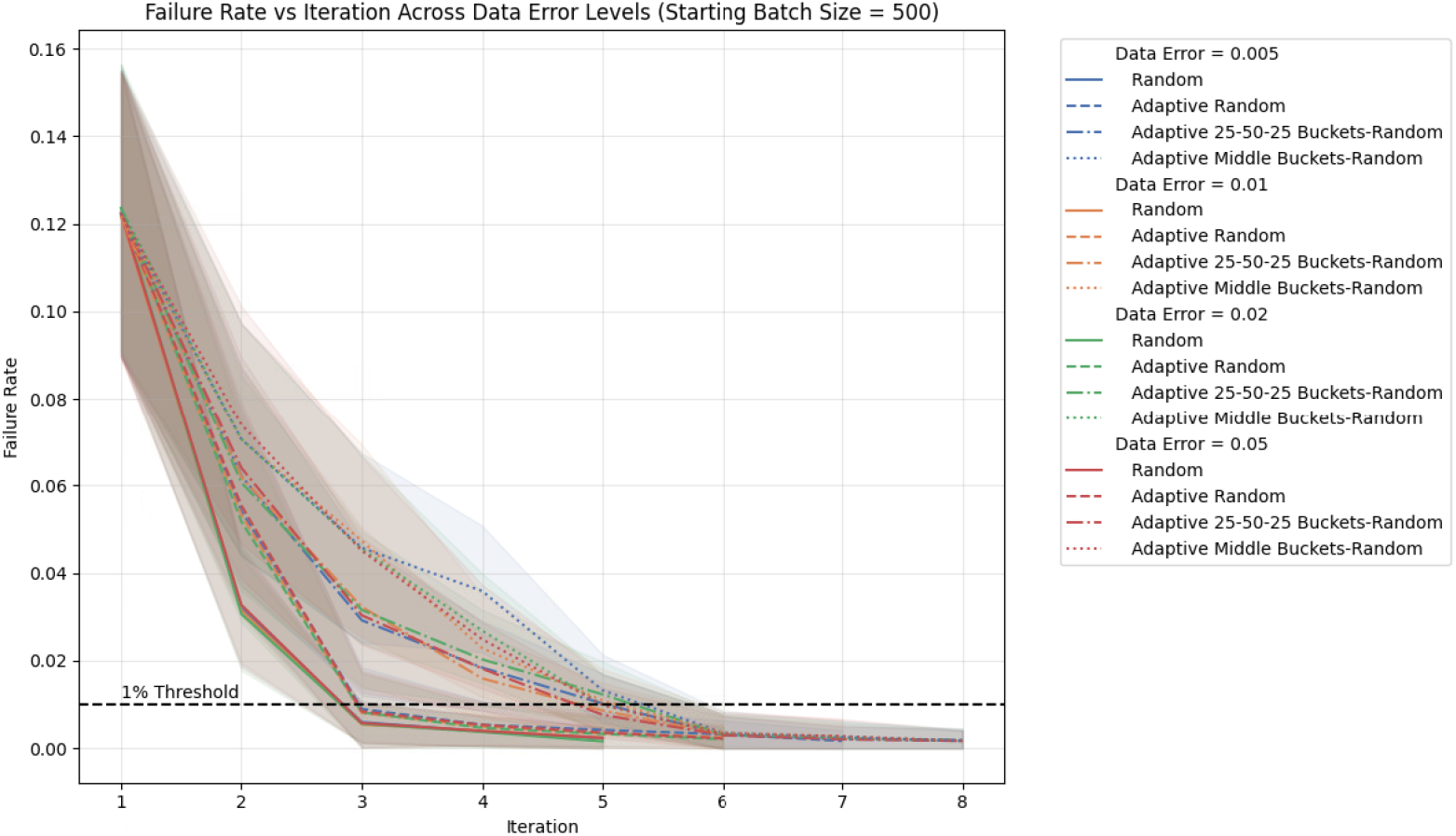
Failure Rate Trajectories Under Varying Data-Error Levels (N = 500 starting batch size)

Figure 5 shows two distinct performance clusters. The Random and Adaptive Random exhibit the steepest early declines and consistently reach the convergence threshold in the fewest iterations. In contrast, the bucket-based strategies, Adaptive Middle Buckets–Random and Adaptive 25-50-25 Buckets–Random, descend more gradually and require more refinement cycles before crossing the threshold. Nevertheless, all strategies ultimately converge below the 1.0% threshold for all data error levels.

Next, we examine the impact of errors in a researcher’s accuracy in reviewing cases. Figure 6 below shows the failure-rate trajectories under simulated reviewer-error levels of 0.5%, 1.0%, 2.0%, and 5.0%. At the lower reviewer-error settings (0.5% and 1.0%), failure rates decline steadily across iterations, and the 95% confidence upper bound approaches or crosses the 1.0% certification threshold for lower error levels. At higher reviewer-error levels, the trajectories continue to decrease but stabilize at values that correspond to the injected reviewer-error rate. For example, under the 5.0% condition, both the failure rate and upper bounds remain near 5% throughout the refinement process.

**Figure 6.**
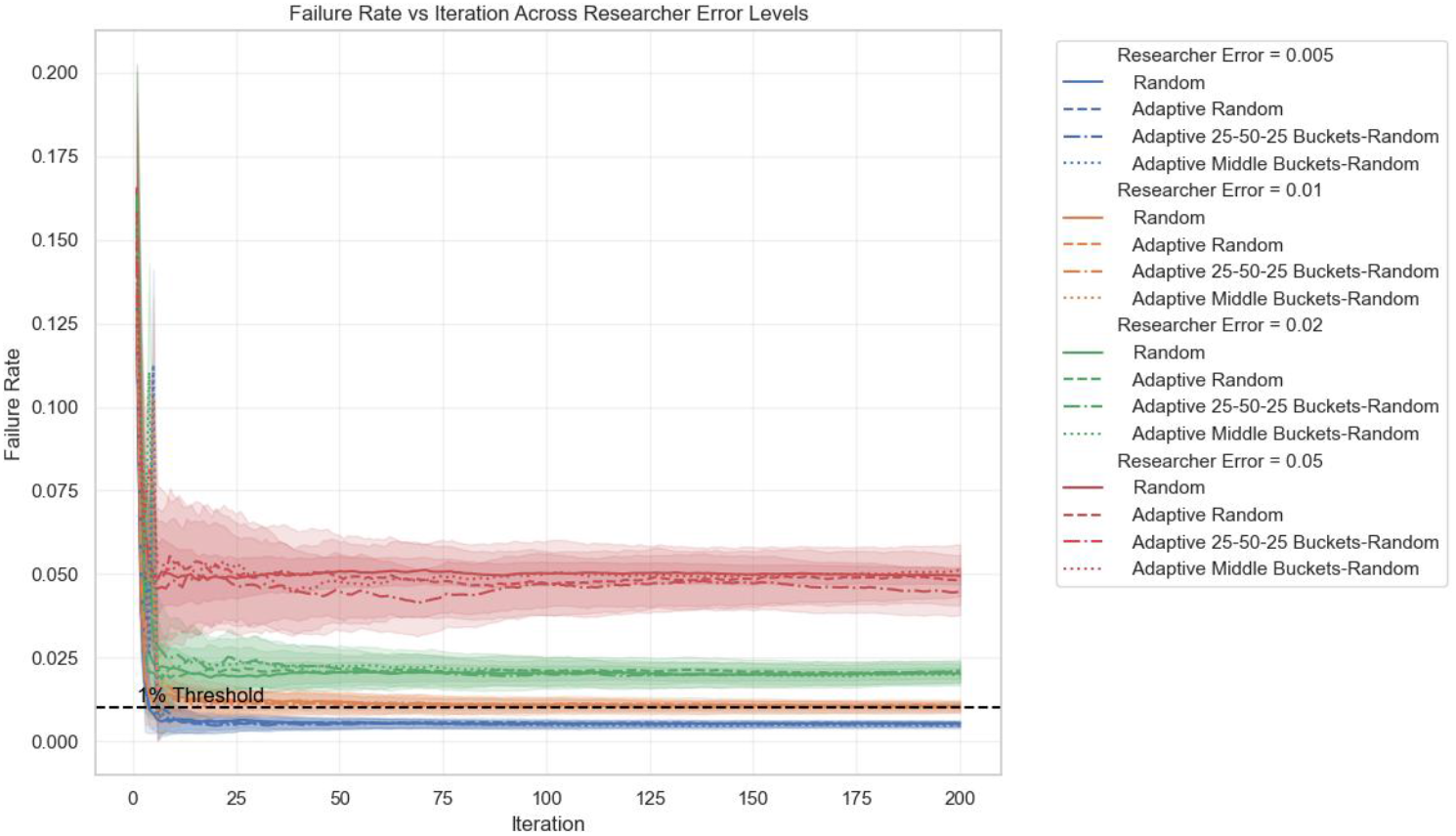
Failure Rate Trajectories Under Varying Researcher-Error Levels

## 4 Discussion

To the best of our knowledge, this is the first study to evaluate a fully integrated framework that combines adaptive sampling with sequential statistical testing to deliver a formal, quantifiable quality guarantee for EHR cohorts. Prior work has largely focused on improving initial phenotype definitions or optimizing extraction logic, whereas the review phase has remained ad hoc and without an accompanying statistical metric. By integrating both data error and human reviewer error along with the evaluation of the framework, this study demonstrates that the framework can operate under realistic sources of uncertainty and provides a mathematically defined stopping point for cohort construction.

This study yields three central findings. First, the framework directly measures cohort accuracy, producing an empirical estimate of the error rate and its uncertainty at each iteration. Second, the framework converges even when the underlying data contains noise and errors, successfully producing intended cohorts when human review is precise. Third, the framework cannot converge to an error threshold lower than the reviewer’s error rate. Across all strategies, the results show that the trade-off between iteration count and total manual review burden is driven by how each method allocates its sampling effort. Fixed strategies reduce the number of iterations when the starting size is large, but they achieve this by front-loading review volume. This pattern explains why starting sizes of 1000 or 2000 converge quickly yet require thousands of cases. Bucket-based fixed methods intensify this effect because they oversample regions with high uncertainty, which causes review volume to increase sharply at larger starting sizes.

Adaptive strategies reduce total review burden by adjusting batch size in response to observed failure rates. This adjustment prevents unnecessary review once the estimate stabilizes. As a result, Adaptive Random and the adaptive bucket-based methods consistently required far fewer total cases while maintaining similar or lower iteration counts. Among these approaches, Adaptive Middle Buckets–Random was particularly efficient because it concentrates sampling in the probability region that is most informative for refining the decision boundary. This focus avoids the large and uninformative batches produced by the fixed counterparts.

### 4.1 Operational Implications of Sampling Strategies

Table 1 and Figure 2 show that iteration count and total cases reviewed vary substantially across selection methods and starting batch sizes. These descriptive patterns demonstrate that the apparent efficiency of a strategy depends on which metric is prioritized. Some methods complete the refinement process in relatively few iterations, while others achieve comparable or lower total review volume. These contrasts introduce the more detailed patterns in Figures 3 and 4, which illustrate how iteration counts and total review burden diverge across methods.

There is no universally optimal sampling strategy because the optimal choice depends on the relative cost of additional iterations compared with the cost of reviewing more cases. Our results show a consistent inverse relationship across all selection methods. Strategies that converge in fewer iterations often require a larger review burden, while strategies that reduce the number of cases reviewed typically involve more refinement cycles. Once a relative cost structure is defined, the framework quantifies these trade-offs and identifies the strategy that is optimal for that specific operational context. This includes settings where turnaround time, reviewer capacity, or tolerance for repeated manual review is the dominant constraint. These findings establish the baseline for the error-injection experiments that follow and describe system behavior under ideal conditions before errors from data quality or reviewer reliability are introduced.

### 4.2 Trajectories in Error-Injected Scenarios

Figure 5 shows that selection strategies respond differently to data errors because each method interacts with the uncertainty structure of the model in distinct ways. Across all error levels, the strategies separate into two consistent behavioral groups. Random and Adaptive Random produce the steepest early declines in failure rate, while the bucket-based strategies reduce failure more gradually. This pattern reflects how each method allocates sampling across the model’s probability distribution. Random and Adaptive Random distribute selections broadly, which allows the system to correct widespread errors quickly. In contrast, the bucket-based strategies focus on intermediate-probability cases, which slows the initial rate of decline but ultimately leads to the same convergence point.

The faster early decline of the Random strategies reflects their broad sampling behavior rather than greater efficiency. These methods expose the model to a representative mix of cases, which allows the framework to correct high-certainty and low-certainty errors at the same time. In contrast, the bucket-based strategies emphasize the uncertain region, which becomes informative only after the model has accumulated enough labeled cases to reliably identify borderline examples. This difference in sampling emphasis explains why the two groups maintain distinct trajectories across all error levels. The proportional scaling of these trajectories across injected-error conditions further shows that the framework responds predictably to changes in baseline data quality. Higher data error shifts the curves upward but does not alter their overall shape. This stability indicates that the system’s corrective logic is robust to varying levels of data error and that strategy choice primarily affects the pace of convergence rather than the final outcome.

Figure 6 shows that reviewer error imposes a hard limit on achievable accuracy because the framework cannot correct mistakes introduced during manual adjudication. In these simulations, the failure rate never drops below the reviewer-error rate, regardless of the selection strategy or the number of iterations. This pattern reflects a structural constraint of the system. The framework can overcome substantial data noise and still converge to the pre-specified threshold, but it cannot compensate for errors made during review. The reviewer’s reliability therefore determines the lowest attainable failure rate.

In real-world settings, these trajectory signatures function as diagnostic indicators of reviewer-driven bottlenecks. When the curves stabilize above the threshold, additional refinement is not the limiting factor; reviewer accuracy is. In such cases, improvements in reviewer training, calibration, or adjudication support are required before further progress is possible. This behavior contrasts with the trajectories in Figure 5, where the system continues to refine the cohort even under substantial data noise. Together, these patterns clarify when refinement remains productive and when the underlying review process must be strengthened to achieve the desired level of accuracy.

### 4.3 Positioning Within Existing Literature

The structure of this framework aligns with longstanding concerns about how EHR cohorts are defined, validated, and reproduced across studies. Existing approaches to cohort construction, whether based on expert-defined rules, phenotype libraries, or site-specific heuristics, often rely on ad hoc review, limited documentation of intended populations, and inconsistent validation practices. The consequences of these gaps are evident in the work of Ostropolets et al.^13^, where nine independent research teams attempting to reproduce the same observational study using the same data produced markedly different cohorts. This level of discordance illustrates how easily hidden assumptions and operational ambiguities can propagate into substantial analytic variability. Because initial cohort definitions influence all downstream analyses, there is a critical need for systematic cohort verification to identify quality issues before the analysis phase. The proposed framework addresses this gap by providing an iterative, data-driven process that refines the cohort definition. By making each refinement step explicit, the framework reduces the likelihood that independent teams will operationalize the same phenotype differently. If our framework were implemented in a similar multi-team setting, it may help surface discrepancies arising from ambiguous criteria, hidden logic, or site-specific interpretations earlier in the workflow, which in turn could support greater alignment across independently constructed cohorts. By shifting from a linear extraction process to the iterative approach described by Jones et al.^21^, the framework also addresses key bottlenecks, including workflow complexity and the substantial labor demands that have historically limited the scalability of gold-standard manual chart review.

### 4.4 Limitations

Several limitations of this study should be noted. First, the rule-revision logic of the simulated researcher is currently fixed, selecting the next rule solely based on the greatest mathematical reduction in error. This design does not fully capture the heuristic decision-making of human researchers, whose choices may incorporate clinical nuance, contextual cues, or qualitative observations and could allow cohort definitions to improve more quickly. Second, the evaluation introduced only random errors into the data and did not model systematic error, such as consistent misclassification patterns or documentation biases that arise from institutional workflows. As a result, the simulation may not fully reflect scenarios in which the data-generation process itself is biased. Third, our simulation of reviewer error used a static error rate. In practice, manual chart review is susceptible to fatigue, learning effects, and phenotype-specific variability, all of which may cause reviewer accuracy to fluctuate over time. Finally, the evaluation was limited to a single clinical condition and a single gold-standard proxy. Although this provides a controlled environment for testing, it may not capture the full variability of diverse clinical phenotypes. For model-based case selection methods, we also relied exclusively on a logistic regression model, which may behave differently from other predictive models.

### 4.5 Future work

Future work should extend this framework to additional error structures, including systematic and other non-random patterns, and evaluate performance across a broader range of clinical conditions and concurrent noise scenarios. Exploring alternative predictive models beyond logistic regression and incorporating diverse gold standards will help assess the generalizability of the approach. In addition, integrating dynamic representations of human behavior, such as fatigue-related variability or expertise-driven decision logic, would allow for a more realistic characterization of reviewer behavior during the manual review process. These future work would position the framework not only as a validation method but also as a foundational component for constructing reproducible and reliable EHR cohorts.

Beyond methodological extensions, the empirical differences among selection strategies suggest an additional opportunity. Because the choice of strategy reflects practical constraints such as available reviewer time, tolerance for iteration, and the need for rapid convergence, future work could formalize how these operational preferences map to specific sampling choices. As more empirical evidence accumulates, it may become possible to identify combinations of strategy and starting batch size that minimize total review burden while preserving statistical certainty. This would allow researchers to tailor the process based on their resource and timeline constraints.

## 5 Conclusion

This study demonstrates that an adaptive, statistically grounded framework can provide a formal quality guarantee for EHR cohort construction. This contribution addresses a longstanding gap in the literature, which has lacked standardized and reproducible methods for assessing the fitness of a constructed cohort before downstream analysis. By integrating adaptive sampling with sequential statistical testing, the framework offers a transparent mechanism for quantifying cohort accuracy, correcting noisy initial definitions, and identifying when reviewer precision is insufficient to meet a desired threshold. The evaluation further shows that operational efficiency is shaped by a generalizable trade-off between iteration count and total review volume, and that adaptive strategies consistently reduce unnecessary review effort. Together, these findings position the framework as both a practical tool for cohort refinement and a diagnostic of cohort quality, thereby improving the rigor, reproducibility, and scalability of EHR-based cohort construction.

## Data Availability

All data produced in the present study are available upon reasonable request to the authors.

